# Occupation and COVID-19 mortality in England: a national linked data study of 14.3 million adults

**DOI:** 10.1101/2021.05.12.21257123

**Authors:** Vahé Nafilyan, Piotr Pawelek, Dan Ayoubkhani, Sarah Rhodes, Lucy Pembrey, Melissa Matz, Michel P Coleman, Claudia Allemani, Ben Windsor-Shellard, Martie van Tongeren, Neil Pearce

## Abstract

**Objective:** To estimate occupational differences in COVID-19 mortality, and test whether these are confounded by factors, such as regional differences, ethnicity and education or due to non-workplace factors, such as deprivation or pre-pandemic health.

**Design:** Retrospective cohort study

**Setting:** People living in private households England

**Participants:** 14,295,900 people aged 40-64 years (mean age 52 years, 51% female) who were alive on 24 January 2020, living in private households in England in 2019, were employed in 2011, and completed the 2011 census.

**Main outcome measures:** COVID-19 related death, assessed between 24 January 2020 and 28 December 2020. We estimated age-standardised mortality rates per 100,000 person-years at risk (ASMR) stratified by sex and occupations. To estimate the effect of occupation due to work-related exposures, we used Cox proportional hazard models to adjust for confounding (region, ethnicity, education), as well as non-workplace factors that are related to occupation.

**Results:** There is wide variation between occupations in COVID-19 mortality. Several occupations, particularly those involving contact with patients or the public, show three-fold or four-fold risks. These elevated risks were greatly attenuated after adjustment for confounding and mediating non-workplace factors. For example, the hazard ratio (HR) for men working as taxi and cab drivers or chauffeurs changed from 4.60 [95%CI 3.62-5.84] to 1.47 [1.14-1.89] after adjustment. More generally, the overall HR for men working in essential occupations compared with men in non-essential occupations changed from 1.45 [1.34 - 1.56] to 1.22 [1.13 - 1.32] after adjustment. For most occupations, confounding and other mediating factors explained about 70% to 80% of the age-adjusted hazard ratios.

**Conclusions:** Working conditions are likely to play a role in COVID-19 mortality, particularly in occupations involving contact with COVID-19 patients or the public. However, there is also a substantial contribution from non-workplace factors, including regional factors, socio-demographic factors, and pre-pandemic health.

## Introduction

The coronavirus pandemic has been particularly severe in the United Kingdom, where high infection and death rates have been reported. Whilst most deaths occur amongst elderly adults [1], many deaths have also occurred among those of working age, particularly among essential workers [2].

Several studies have reported important occupational differences in the risk of SARS-CoV-2 infection and death [3, 4, 5], but there have been relatively few systematic comparisons of death rates in different occupations. Infections in health care workers have received the most attention [6, 7], with evidence that intensive care unit workers who care for COVID-19 patients are at elevated risk. However, other occupations may also be at increased risk, particularly those which involve social care, or contact with the public [8]. In particular, age-standardised mortality rates (ASMRs) for COVID-19 by occupation are high among taxi drivers and chauffeurs, bus and coach drivers, chefs, sales and retail assistants, and social care workers [9].

Occupational inequality in COVID-19 mortality is a major public health problem [10, 8], but it is challenging to determine the extent to which working conditions drive these raised risks. Occupational differences in COVID-19 mortality could be caused by non-workplace factors such as living conditions at home or poor underlying (pre-pandemic) health. Deprivation, poor health and occupation are all linked. For example, people working in low-paid, insecure jobs are also likely to experience poor housing conditions, overcrowding and low pre-pandemic health status. COVID-19 mortality is also higher in people with specific comorbidities [11, 12]. However, it is important to distinguish between situations where high COVID-19 mortality rates in a particular occupational group are directly due to workplace exposures, and those which are due to non-workplace factors. This distinction is particularly important for public health policy. If the excess risk in an occupation (e.g. bus drivers) is due to working conditions, this would suggest the need for preventive interventions in the workplace. However, if the excess is due to non-workplace factors such as living conditions at home (which may be associated with working conditions, but are not caused by them), then different interventions would be required. In both instances, the observed differences in risk would be real, and preventable, but the policy implications would be different.

In this study, we estimated occupational differences in COVID-19 mortality in England and Wales during 2020. We have examined how much these differences changed after adjustment for non-workplace factors, using Cox proportional hazard models.

## Methods

### Data

We used individual-level data from the Public Health Data Asset. This dataset is based on the 2011 Census in England, linked with the NHS number to death records, Hospital Episode Statistics and the General Practice Extraction Service (GPES) data for pandemic planning and research. To obtain NHS numbers, the 2011 Census was linked to the 2011-2013 NHS Patient Registers, using deterministic and probabilistic matching, with an overall linkage success of 94.6%. We excluded individuals (12.4%) who did not have a valid NHS number or were not linked to GPES primary care records. We used data on 14,295,900 individuals who were aged 31-55 years at the time of the 2011 Census and were therefore likely to be in stable employment both in 2011 and 2020 (by which time they were aged 40-64 years). We examined the differences between occupation groups in the risk of death involving COVID-19 during the 11 months from 24 January to 28 December 2020.

### Outcomes

Individuals in the study population were followed up from 24 January until 28 December 2020 for COVID-19 death (either in hospital or out of hospital), defined as confirmed or suspected COVID-19 death as identified by one of two ICD10 (International Classification of Diseases, 10th revision) codes (U07.1 or U07.2) derived from the medical certificate of cause of death.

### Exposure

We chose the main exposure as the occupation at the time of the 2011 Census. Occupations are coded using a hierarchical classification, under the Standard Occupation Classification (SOC) 2010 (7). The most detailed classification (Unit group, with 4-digit codes) includes 369 categories, whilst the most aggregated (Major group, with 1-digit codes) has only nine groups.

We derived a hybrid classification based on the sub-major groups (2-digit codes), which include 25 categories. We also examined some 3-digit and 4-digit codes to assess selected occupations that have previously been shown to have high COVID-19 mortality, such as taxi drivers, security guards, or care home workers [4]. Our final classification contained 41 categories (Supplementary Table S1 in Appendix). We derived a classification of essential workers, based on the classification developed for a recent study using data from the UK Biobank [3].

Because we used the occupation recorded at the 2011 Census, our exposure variable is likely to be misclassified for some participants, since people may have left the labour force or changed occupation since 2011. To estimate the extent of misclassification, we used data from Understanding Society, a large-scale longitudinal household survey, to analyse occupational mobility across Major (1-digit SOC codes) groups between 2011 and 2019.

### Covariates

We aimed to distinguish between situations where high COVID-19 mortality rates in a particular occupational group are likely to be directly due to work-place exposures, and those that may be due to non-workplace factors. In addition to the basic age-adjusted models, we adjusted for potential confounders such as geography and ethnicity. We also adjusted for factors that may be related to occupation, but which affect exposures outside the workplace, including markers of deprivation and housing conditions. These are all potential confounders of the association between workplace exposures and COVID-19, because they may be associated with the risk of COVID-19 mortality, either through the propensity to become infected or the propensity to die once infected. All covariates are summarised in Supplementary Table S2 in the Appendix. Geographical factors and socio-demographic characteristics were based on the 2011 Census; body mass index (BMI) and comorbidities were derived from the primary care and hospitalisation data following the definitions adopted by the QCOVID risk prediction model [11].

### Statistical analyses

For the period from 24^th^ January 2020 to 28^th^ December 2020, we calculated age-standardized mortality rates (ASMRs) for each occupation using the European Standard Population [13].

To estimate the effect of occupation due to work-related exposures, we used Cox proportional hazard models to adjust for confounding (region, ethnicity, education), as well as non-workplace factors that are related to occupation. We estimated five models, sequentially adjusting for additional covariables to assess how they might confound or mediate differences in workplace exposure on the risk of death from COVID-19 (See Figure 1). Our first model was only adjusted for age. The second model also adjusted for geographical factors (region, population density, rural urban classification) to account for the differential spread of the virus in different areas. The third model further adjusted for other confounding factors, ethnicity and education, which are related both to occupation and COVID-19 risk. The fourth model also controlled for non-workplace factors (living conditions), including socio-economic factors (Index of Multiple Deprivation, household deprivation, household tenancy and house type) and household composition (household size, children in the household, overcrowding). Finally, the last model adjusted for pre-pandemic health (BMI, chronic kidney disease, learning disability, cancer or immunosuppression, and other conditions; see Supplementary Table S2 for details on all the covariates). We used corporate managers and directors as the reference category, because it is a large group with a low absolute risk [9].

**Figure 1.**
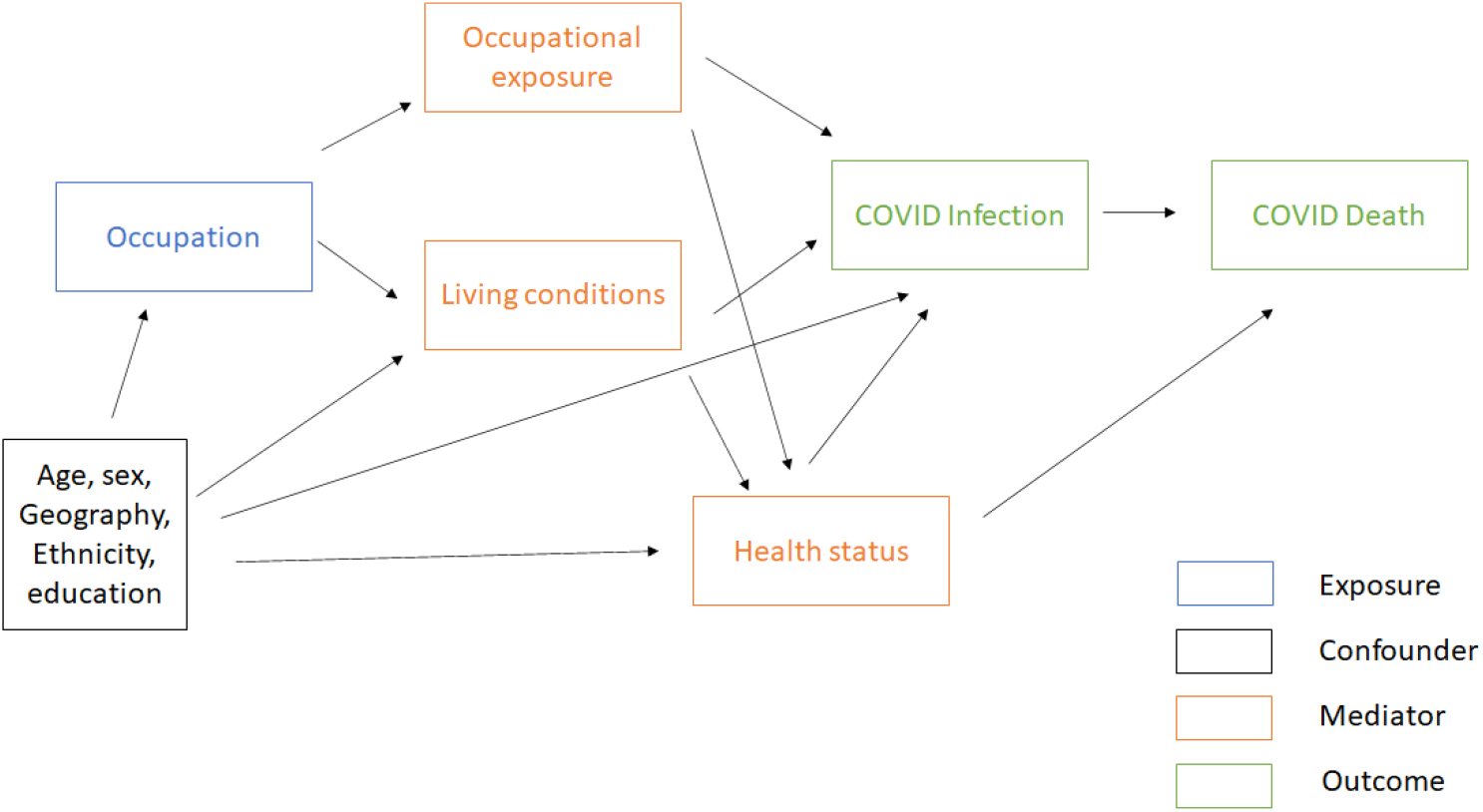
Directed acyclic graph of the relationship between occupation and COVID-19 mortality.

## Results

### Characteristics of the study population

Our analytical sample comprises 14,295,900 people aged 40-64 years (mean age 52 years, 51% female) who were alive on 24 January 2020, living in private households in England in 2019, were employed in 2011, and completed the 2011 census. Between 24 January and 28 December 2020, 4,552 people (0.003%) died from a cause related to COVID-19; characteristics of these individuals are summarized in Table 1 (further details in Supplementary Table S3).

**Table 1.**
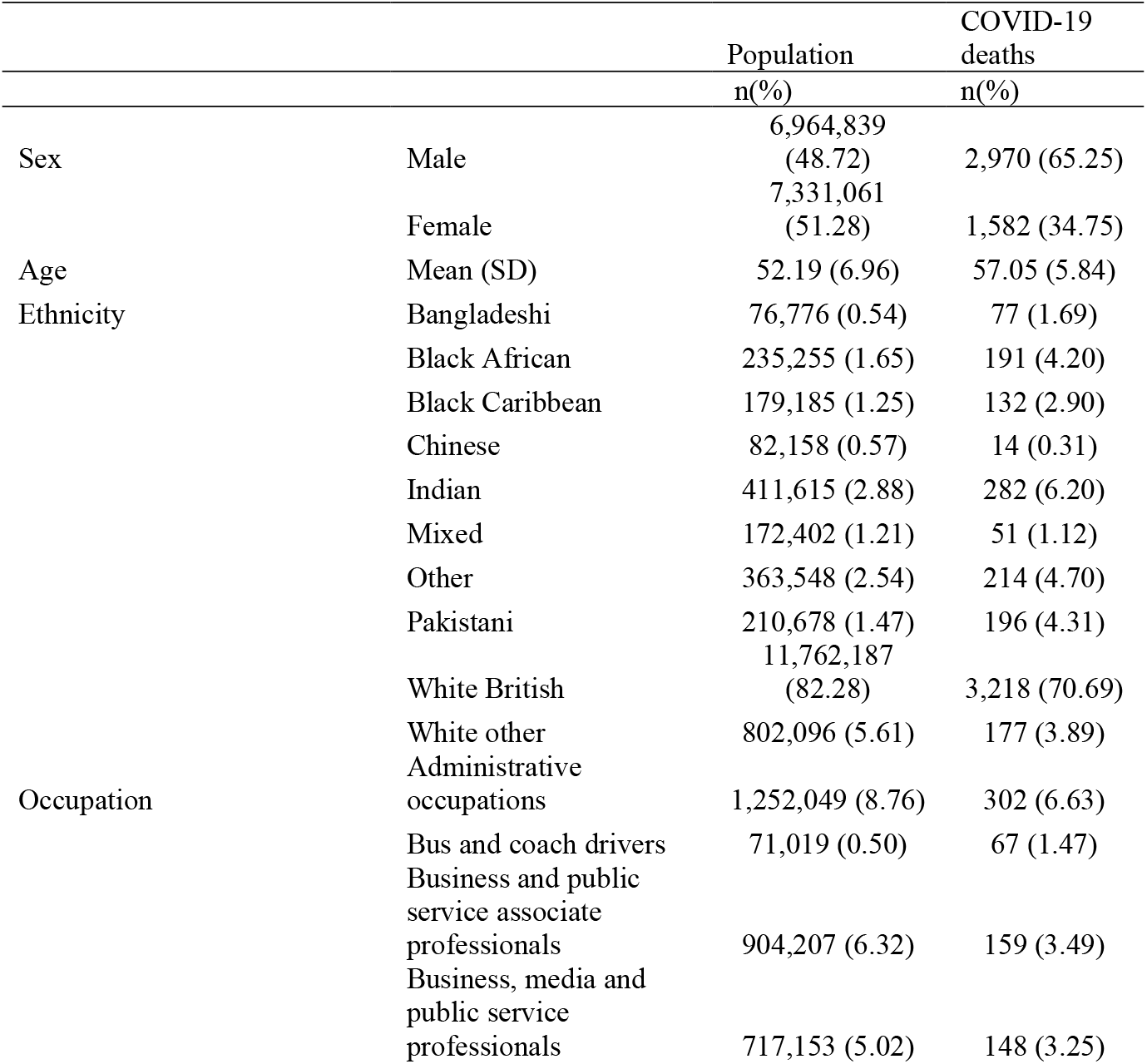
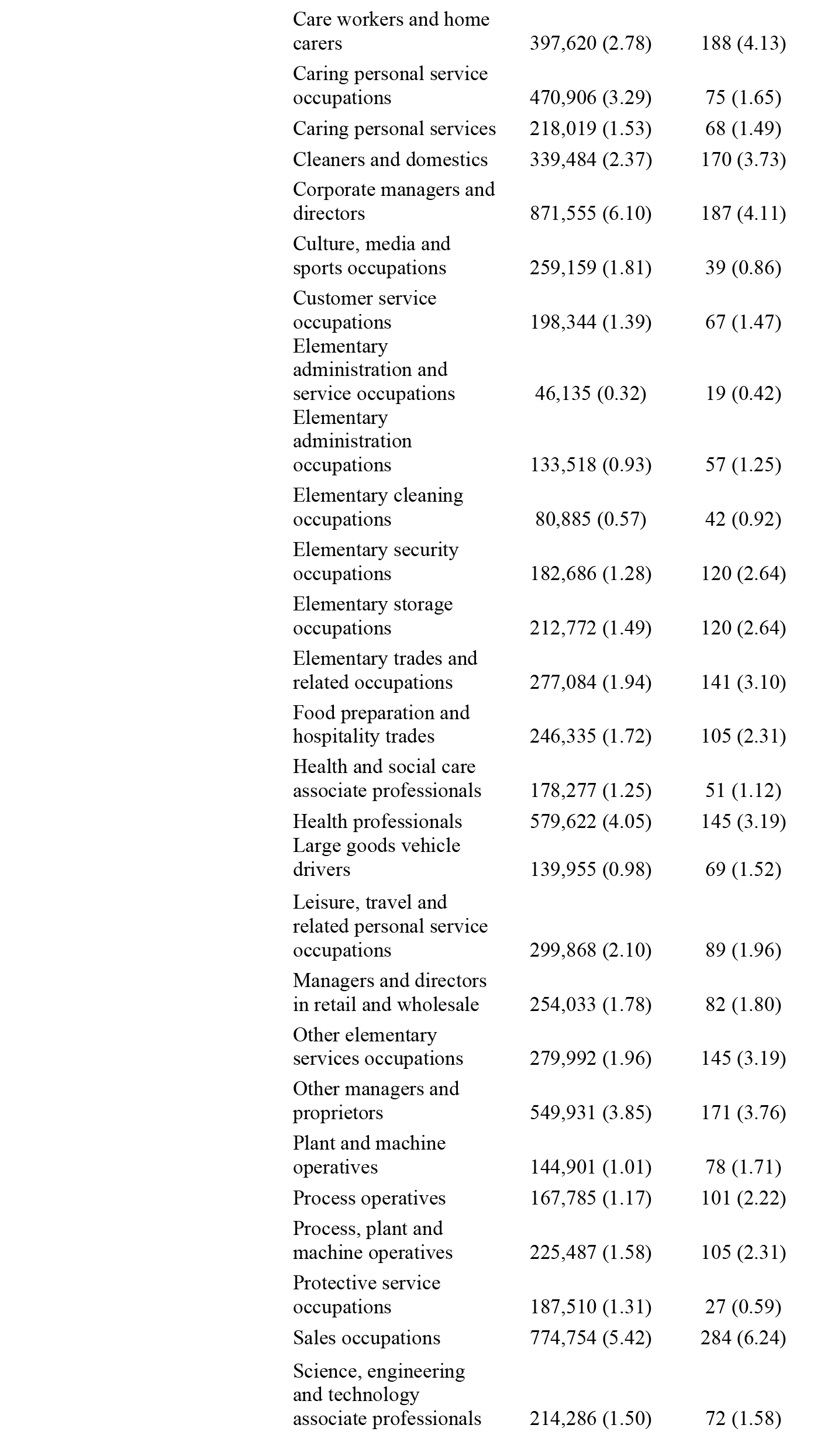
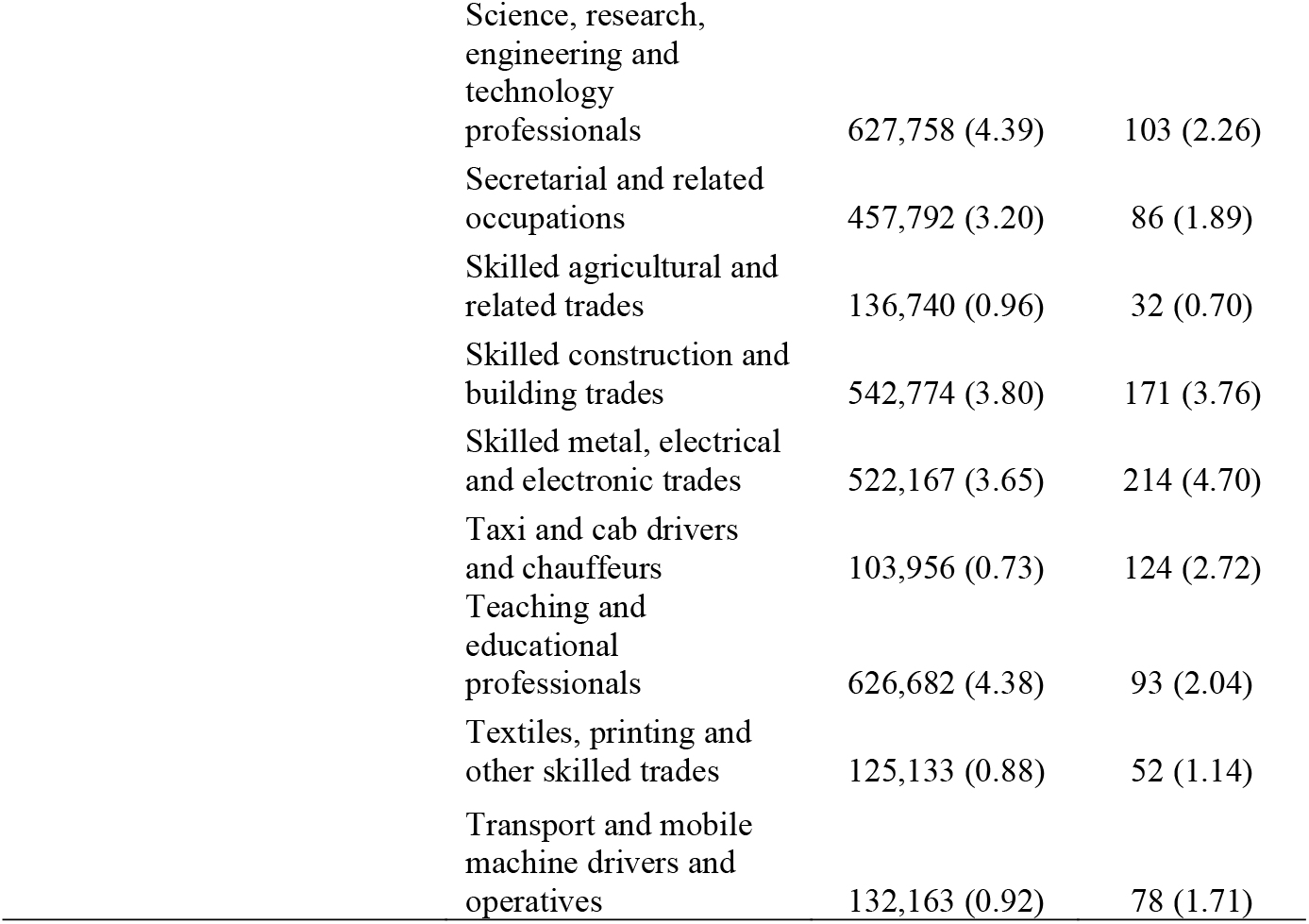
Characteristics of the study population and those who died from a cause related to COVID-19.

### Age-standardized mortality rates

Table 2 shows the annualised ASMRs for Covid-19 for men aged 40 to 64 years. The ASMRs were highest among those working as taxi and cab drivers or chauffeurs at 119.7 deaths per 100,000 men [95% Confidence Interval: 98.0 - 141.4] over the period, followed by other elementary occupations at 106.5 [84.5 – 132.4] and care workers and home carers at 99.2 [74.5 – 129.4] (Table 2). The ASMRs were lowest among those working in the protective service occupations at 19.5 [12.5 - 28.8], followed by science, research, engineering and technology professionals at 20.9 [16.8 - 25.6] and skilled agricultural and related trades occupations at 23.4 deaths per 100,000 men [15.3 - 34.0].

**Table 2.** Annualised Age-standardised mortality rates per 100,000 adults aged 40-64 years, by sex and occupation.

| **Occupation** | **Men** | **Women** |
| --- | --- | --- |
| Taxi and cab drivers and chauffeurs | 119.7 [98.0 - 141.4] | - |
| Other elementary occupations | 106.5 [84.5 - 132.4] | 36.5 [29.1 - 45.3] |
| Care workers and home carers | 99.2 [74.5 - 129.4] | 39.4 [32.7 - 46.1] |
| Secretarial and related occupations | 96.2 [57.0 - 152.0] | 15.6 [12.0 - 19.9] |
| Elementary security occupations | 93.8 [74.7 - 116.2] | 40.5 [28.3 - 56.2] |
| Bus and coach drivers | 90.8 [69.0 - 117.2] | - |
| Cleaners and domestics | 84.2 [62.2 - 111.4] | 40.0 [32.7 - 47.3] |
| Customer service occupations | 75.2 [52.2 - 104.8] | 25.3 [17.3 - 35.7] |
| Van drivers | 71.6 [57.5 - 85.8] | - |
| Caring personal services | 69.1 [45.0 - 101.3] | 23.0 [16.5 - 31.2] |
| Process operatives | 65.2 [51.2 - 81.8] | 46.9 [30.3 - 69.2] |
| Food preparation and hospitality trades | 64.3 [50.4 - 81.0] | 28.2 [19.0 - 40.1] |
| Sales occupations | 63.8 [51.7 - 75.9] | 30.4 [25.9 - 34.9] |
| Elementary storage occupations | 63.5 [51.4 - 75.6] | 37.8 [20.6 - 63.4] |
| Mobile machine and other drivers | 59.3 [46.4 - 74.5] | - |
| Health and social care associate professionals | 59.2 [38.2 - 87.5] | 20.1 [13.1 - 29.4] |
| Health professionals | 54.2 [41.7 - 69.2] | 18.7 [14.8 - 23.2] |
| Elementary trades and related occupations | 53.3 [43.2 - 64.9] | 45.2 [32.4 - 61.3] |
| Leisure, travel and related personal service occupations | 53.2 [38.5 - 71.7] | 22.8 [16.7 - 30.5] |
| Elementary administration occupations | 52.0 [38.2 - 69.3] | - |
| Administrative occupations | 51.2 [42.6 - 59.8] | 17.0 [14.4 - 19.6] |
| Elementary cleaning occupations excl. cleaners and domestics | 48.9 [32.7 - 70.1] | 51.7 [27.5 - 88.4] |
| Textiles, printing and other (excl. food prep and hospitality) skilled trades | 48.8 [35.0 - 66.2] | 23.0 [11.4 - 41.2] |
| Assemblers and construction operatives | 48.2 [37.9 - 60.4] | 36.7 [24.0 - 53.2] |
| Large goods vehicle drivers | 47.3 [36.3 - 60.5] | - |
| Plant and machine operatives | 45.3 [34.6 - 58.2] | 77.2 [43.1 - 126.8] |
| Science, engineering and technology associate professionals | 44.3 [33.8 - 57.1] | 20.1 [10.4 - 35.2] |
| Managers and directors in retail and wholesale | 43.9 [33.4 - 56.7] | 23.6 [15.0 - 35.5] |
| Skilled metal, electrical and electronic trades | 40.4 [34.9 - 45.9] | - |
| Other managers and proprietors | 37.9 [31.0 - 44.8] | 20.5 [15.3 - 26.9] |
| Caring personal service occupations excl. care workers and home carers | - | 15.4 [11.9 - 19.7] |
| Skilled construction and building trades | 31.5 [26.7 - 36.2] | - |
| Business, media and public service professionals | 27.3 [22.0 - 32.6] | 17.2 [12.6 - 23.1] |
| Teaching and educational professionals | 26.0 [18.9 - 34.8] | 11.2 [ 8.3 - 14.9] |
| Culture, media and sports occupations | 25.8 [17.3 - 36.8] | - |
| Corporate managers and directors excl. those in retail and wholesale | 25.6 [21.5 - 29.7] | 12.5 [ 8.5 - 17.7] |
| Business and public service associate professionals | 25.6 [20.8 - 30.4] | 14.0 [10.4 - 18.6] |
| Skilled agricultural and related trades | 23.4 [15.3 - 34.0] | - |
| Science, research, engineering and technology professionals | 20.9 [16.8 - 25.6] | 10.3 [ 5.0 - 18.7] |
| Protective service occupations | 19.5 [12.5 - 28.8] | - |

Note: Deaths occurring between 24 January 2020 and 28 December 2020. 95% confidence intervals are reported. Mortality rates are standardised to the 2013 European Standard Population and annualised. Occupation classification and associated SOC 2010 codes are given in Supplementary Table S1. Data are not reported if there were 10 or fewer deaths

For women aged 40 to 64 years, the ASMRs for COVID-19 mortality were greatest among those working as process operatives at 46.9 deaths per 100,000 women [30.3 - 69.2], followed by elementary trades and related occupations at 45.2 [32.4 - 61.3] and elementary security operations at 40.5 [28.3 - 56.2] (Table 2). The ASMRs were lowest among those working in the teaching and educational professions at 11.2 [8.3 - 14.9], followed by corporate manager and directors, excluding those in retail and wholesale at 12.5 [8.5 - 17.7] and business and public service associate professionals at 17.2 [12.6 - 23.1].

### Adjusted Hazard Ratios

The age-adjusted Hazard ratios (HRs), relative to corporate managers, indicated large differences in COVID-19 mortality between occupations for both men and women (Figure 1). For men, adjustment for confounding factors strongly attenuated the hazard ratios for many occupations, but several remained at elevated risk after adjustment. Men working in secretarial and related occupations remained at elevated risk of dying from COVID-19 with a fully adjusted HR of 1.90 [1.16 - 3.11], compared to an age-adjusted HR of 3.82 [2.34 - 6.22]. Care workers and home carers were also at higher risk of COVID-19 death, with a fully adjusted HR of 1.75 [1.28 - 2.40] compared to an age-adjusted HR of 3.85 [2.83 - 5.25]. Working as taxi and cab drivers or chauffeurs was associated with elevated risk with a fully adjusted HR of 1.47 [1.14 - 1.89], compared to an age-adjusted HR of 4.60 [3.62 - 5.84]. Other occupations with a high risk of COVID-19 death after adjustment for confounding factors included health professionals (1.67 [1.24 - 2.25]), caring personal services (1.67 [1.09 - 2.53]), health and social care associate professionals (1.63 [1.07 - 2.49]), customer service occupation (1.60 [1.10 - 2.32]), other elementary occupations (1.48 [1.11 - 1.95]), administrative occupations (1.33 [1.05 - 1.68]) and bus and coach drivers (1.36 [1.00 - 1.84]).

For women, adjusting for confounding factors also greatly attenuated the estimated difference in risk between occupations. The highest age-adjusted HRs were observed for plant and machine operatives (5.54 [3.04 - 10.10]]) and those working elementary cleaning (5.54 [3.04 - 10.10]), but no occupation groups experienced a significantly elevated risk of COVID-19 mortality after adjusting for other factors. For many occupations, the HRs are of similar magnitude to those for men, but less precise because of smaller numbers.

For most occupations, confounding and other mediating factors explained about 70% to 80% of the age-adjusted hazard ratios. Adjusting for socio-economic status had the largest impact on the hazard ratios, followed by geographical factors (See Supplementary Table S4 and S5 for men and women respectively). A notable exception is health professionals, for whom adjustment for socio-economic factors did not affect the hazard ratios. Hazard ratios obtained when using all other occupations (rather than corporate managers and directors) as a reference group are similar – the unadjusted hazard ratios are slightly lower, but the adjusted estimates are similar to those in our main analyses (Supplementary Table S7)

**Figure 1.**
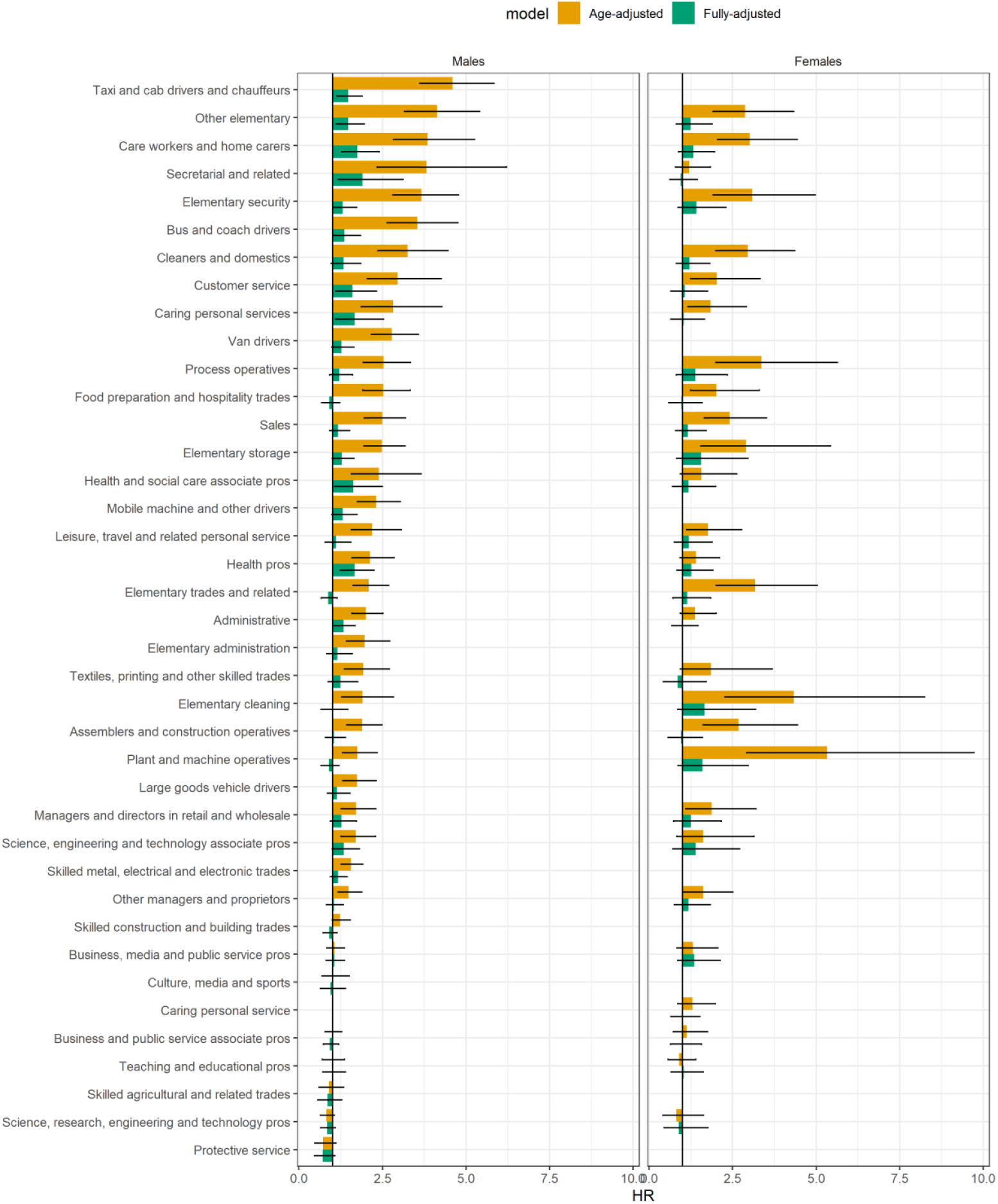
Hazard ratios for COVID-19 related death for adults aged 40 to 64 years, compared to corporate managers and directors, by sex. Note: Fully adjusted Cox regression models include geographical factors (region, population density, urban/rural classification), ethnicity, socio-economic characteristics (Index of Multiple Deprivation decile group, household deprivation, educational attainment, social grade, household tenancy, type of accommodation, household size, multigenerational household, household with children), health (body mass index, chronic kidney disease (CKD), learning disability, cancer and immunosuppression, other conditions). See Supplementary Tables A1 for more details. Occupation classification and associated SOC 2010 codes are given in Supplementary Table S1. Numerical results can be found in Supplementary Tables S4 and S5.

Table 4 shows the hazard ratios for essential workers compared to non-essential workers as the reference category. Overall, essential workers are at higher risk of COVID-19 death than non-essential workers, and most categories of essential workers also have higher mortality. Once again, the differences are generally much attenuated after adjusting for potential confounding and mediating factors; a notable exception is health care professionals. We also report hazard ratios for major groups, compared to directors and managers, in Supplementary Table S7.

**Table 4.** Hazard ratios for COVID-19 related death for adults aged 40 to 64 years, compared to non-essential workers, by sex.

| Occupation | Men |  | Women |  |
| --- | --- | --- | --- | --- |
|  | Age-adjusted | Fully adjusted | Age-adjusted | Fully adjusted |
| Essential workers | 1.45 [1.34 - 1.56] | 1.22 [1.13 - 1.32] | 1.16 [1.05 - 1.28] | 1.06 [0.96 - 1.17] |
| Taxi and cab drivers and chauffeurs | 3.08 [2.56 - 3.70] | 1.39 [1.14 - 1.70] | 3.94 [1.634 - 9.48] | 2.45 [1.014 - 5.92] |
| Support staff | 2.39 [1.68 - 3.41] | 1.74 [1.22 - 2.49] | 0.95 [0.673 - 1.34] | 0.78 [0.550 - 1.10] |
| Bus and coach drivers | 2.33 [1.81 - 3.00] | 1.11 [0.85 - 1.45] | 2.95 [1.226 - 7.12] | 1.73 [0.716 - 4.18] |
| Sanitary workers | 1.84 [1.46 - 2.32] | 1.18 [0.93 - 1.50] | 1.78 [1.473 - 2.16] | 1.09 [0.892 - 1.33] |
| Social care | 1.83 [1.51 - 2.20] | 1.27 [1.04 - 1.53] | 1.62 [1.390 - 1.89] | 1.18 [1.010 - 1.39] |
| Van drivers | 1.81 [1.48 - 2.22] | 1.26 [1.03 - 1.55] | 1.59 [0.661 - 3.84] | 1.27 [0.526 - 3.06] |
| Health associate professionals | 1.65 [1.26 - 2.16] | 1.86 [1.41 - 2.46] | 0.92 [0.746 - 1.15] | 1.22 [0.969 - 1.54] |
| Food retail & distribution | 1.41 [1.22 - 1.63] | 1.14 [0.98 - 1.32] | 1.39 [1.187 - 1.63] | 1.02 [0.867 - 1.20] |
| Other transport workers | 1.21 [1.02 - 1.43] | 1.10 [0.93 - 1.30] | 0.36 [0.115 - 1.11] | 0.31 [0.098 - 0.95] |
| Health professionals | 1.21 [0.82 - 1.78] | 1.45 [0.97 - 2.15] | 0.25 [0.079 - 0.76] | 0.45 [0.145 - 1.42] |
| Food production | 1.12 [0.86 - 1.45] | 1.15 [0.89 - 1.50] | 1.48 [0.968 - 2.26] | 1.15 [0.750 - 1.77] |
| Education | 0.63 [0.47 - 0.84] | 0.91 [0.68 - 1.23] | 0.56 [0.446 - 0.70] | 0.83 [0.653 - 1.05] |
| Police & Protective Services | 0.45 [0.31 - 0.67] | 0.60 [0.40 - 0.88] | 0.38 [0.123 - 1.19] | 0.50 [0.160 - 1.54] |

Note: Fully adjusted Cox regression models include geographical factors (region, population density, urban/rural classification), ethnicity, socio-economic characteristics (Index of Multiple Deprivation decile group, household deprivation, educational attainment, social grade, household tenancy, type of accommodation, household size, multigenerational household, household with children), health (body mass index, chronic kidney disease (CKD), learning disability, cancer or immunosuppression, other conditions). See Supplementary Tables S1 for more details.

## Discussion

By combining data from the 2011 Census with electronic health records, the Public Health Data Asset has enabled us to analyse detailed information on a wide range of socio-demographic characteristics and individual (pre-pandemic) health status. Information on occupation is not available in traditional electronic health records in the United Kingdom, and the Census is the only source of population-wide occupation data. Our dataset contains over 14 million people aged 40 to 64 years who were living in England at the beginning of the pandemic. We were therefore able to estimate COVID-19 mortality for detailed occupational groups, and to estimate whether the differences in mortality are driven by workplace-related factors, or by other confounding and mediating factors.

The main limitation of our study is that the information on occupation is nine years out of date. Our exposure is therefore likely to be misclassified for a proportion of people, because they have left the labour force or changed occupation since 2011. To mitigate measurement error, we restricted our analysis to people aged 40-64 years, who had a relatively high occupational stability, as shown in our analysis of a large longitudinal household survey. Exposure misclassification is nonetheless likely to result in biasing of the estimated hazard ratios towards the null value of 1.0. However, we still observed strongly elevated hazard ratios for many occupations. Misclassification of occupation would be constant across our various analyses and could not explain the substantial decrease in most hazard ratios after adjustment for confounders. On the other hand, the confounders that we have addressed are also likely to be misclassified to some extent. Given that adjustment for confounders produced large changes in the estimated occupational associations, it is possible that if more accurate or detailed confounder data were available, adjustment would have driven the hazard ratio estimates even lower towards the null value of 1.0.

Another limitation is that our dataset excludes recent migrants, since it is based on people who were enumerated at the 2011 Census. Finally, some deaths may not have been registered by the end of the study period if they had been sent to a coroner, which could affect some occupational groups such as healthcare workers.

Our age-adjusted results are consistent with official estimates of COVID-19 mortality by occupation group [9]. However, we find that these elevated risks were greatly attenuated after adjustment for non-workplace factors, such as geographic factors, socio-demographic factors and pre-pandemic health. A recent study based on the UK Biobank found that compared to non-essential workers, medical support staff and healthcare professionals had the highest risk of severe COVID-19 [3]. We also found that, amongst men, healthcare professionals were at increased risk of death from COVID-19, but the HRs for healthcare professionals in our study were similar to those working as care workers, taxi drivers or in secretarial occupations, after full adjustment. Our results are also consistent with US studies documenting higher mortality rates in essential workers, such as transportation/logistics workers and healthcare workers [5, 14].

Our findings are also generally consistent with a recent analysis of data from the UK Coronavirus Infection Survey, which found increased risks for a similar list of occupations [15]. This analysis found that the occupational differences largely disappeared after adjustment for other factors, but the adjustment included factors that are likely to be inherent to working conditions (inability to work at home, and inability to socially distance at work), and are therefore on the causal pathway linking occupational exposure and infection. Thus, our adjusted findings are not directly comparable with those obtained from the Coronavirus Infection Survey.

Our age-standardised mortality rates and age-adjusted hazard ratios confirm that there is a wide variation in the risk of COVID-19 mortality between occupations. However, workplace exposure is only one of several possible factors that drive the observed differences in the risk of COVID-19 mortality between occupations: other factors also contribute to the observed differences. After adjusting for these factors, people who work in occupations that involve contacts with patients (e.g. health and social care workers) or the public (e.g. bus and taxi drivers, retail workers) remain at elevated risk COVID-19 related death.

Other occupations that do not involve contact with patients or the public may also have increased risks due to specific working conditions (e.g. overcrowding in the workplace, lack of ventilation, lack of PPE, etc), but our analyses indicate that these relative risks are generally small, after adjustment for confounding. This does not mean that infection is not occurring in specific workplaces. Whilst there have been a number of workplace outbreaks reported in various industries such as food processing which do not involve patient or public contact [16, 17], it appears that such outbreaks are not sufficient to produce strongly elevated sector-wide increased risks after adjustment for non-workplace factors.

Our analyses have confirmed previous findings that many occupations have elevated risks of COVID-19 mortality. These associations were greatly attenuated, for many occupations, after adjustment for measures of deprivation and geographical factors, suggesting that differences in risk between occupations are a result of a complex mix of different factors. A number of occupations showed increased risks, even after comprehensive adjustment, and it is likely that working conditions played a role. However, our findings also indicate that non-workplace factors also play a major role. Preventive measures therefore need to reduce workplace exposures, but also to reduce exposures outside the workplace, including overcrowding, inadequate housing, and deprivation.

## Data Availability

The ONS Public Health Linked Data Asset will be made available on the ONS Secure Research Service for Accredited researchers. Researchers can apply for accreditation through the Research Accreditation Service.

## Acknowledgements

We thank Raymond Agius, Sheila Bird, David Coggon, Steve Robertson, and Lesley Rushton for their comments on the draft manuscript.

## Ethics approval

Ethical approval was obtained from the National Statistician’s Data Ethics Advisory Committee (NSDEC(20)12)

## Authors and contributors

Study conceptualisation was led by VN and NP. All authors contributed to the development of the research question, study design, with development of statistical aspects led by VN, NP and PP. VN and PP were involved in data specification, curation and collection. VN and PP conducted and checked the statistical analyses. All authors contributed to the interpretation of the results. VN and NP wrote the first draft of the paper. All authors contributed to the critical revision of the manuscript for important intellectual content and approved the final version of the manuscript.

VN had full access to all data in the study and takes responsibility of the integrity of the data and the accuracy of the data analysis. The lead author (VN) affirms that the manuscript is an honest, accurate, and transparent account of the study being reported; that no important aspects of the study have been omitted; and that any discrepancies from the study as planned have been explained.

## Dissemination

Dissemination of the results to study participants is not possible.

## Patient and Public Involvement

Patients or the public were not involved in the design, or conduct, or reporting, or dissemination plans of our research.

## Appendix

**Supplementary Table S1.** Occupation classification and associated SOC 2010 codes.

| **Occupation** | **SOC**<br>**2010**<br>**code(s)** |
| --- | --- |
| Corporate managers and director (excluding those in retail and wholesale) | 11 |
| Other managers and proprietors | 12 |
| Science, research, engineering and technology professionals | 21 |
| Health professionals | 22 |
| Teaching and educational professionals | 23 |
| Business, media and public service professionals | 24 |
| Science, engineering and technology associate professionals | 31 |
| Health and social care associate professionals | 32 |
| Protective service occupations | 33 |
| Culture, media and sports occupations | 34 |
| Business and public service associate professionals | 35 |
| Administrative occupations | 41 |
| Secretarial and related occupations | 42 |
| Skilled agricultural and related trades | 51 |
| Skilled metal, electrical and electronic trades | 52 |
| Skilled construction and building trades | 53 |
| Textiles, printing and other (excluding food preparation and hospitality) skilled trades | 54 |
| Caring personal service occupations (excluding care workers and home carers) | 61 |
| Leisure, travel and related personal service occupations | 62 |
| Sales occupations | 71 |
| Customer service occupations | 72 |
| Assemblers and construction operatives | 81 |
| Mobile machine and other drivers | 82 |
| Elementary trades and related occupations | 91 |
| Other elementary occupations | 92 |
| Food preparation and hospitality trades | 543 |
| Caring personal services | 614 |
| Process operatives | 811 |
| Plant and machine operatives | 812 |
| Elementary administration occupations | 921 |
| Elementary cleaning occupations (excluding cleaners and domestics) | 923 |
| Elementary security occupations | 924 |
| Other elementary occupations | 927 |
| Managers and directors in retail and wholesale | 1190 |
| Care workers and home carers | 6145 |
| Large goods vehicle drivers | 8211 |
| Van drivers | 8212 |
| Bus and coach drivers | 8213 |
| Taxi and cab drivers and chauffeurs | 8214 |
| Cleaners and domestics | 9233 |
| Elementary storage occupations | 9260 |

**Supplementary Table S2:**
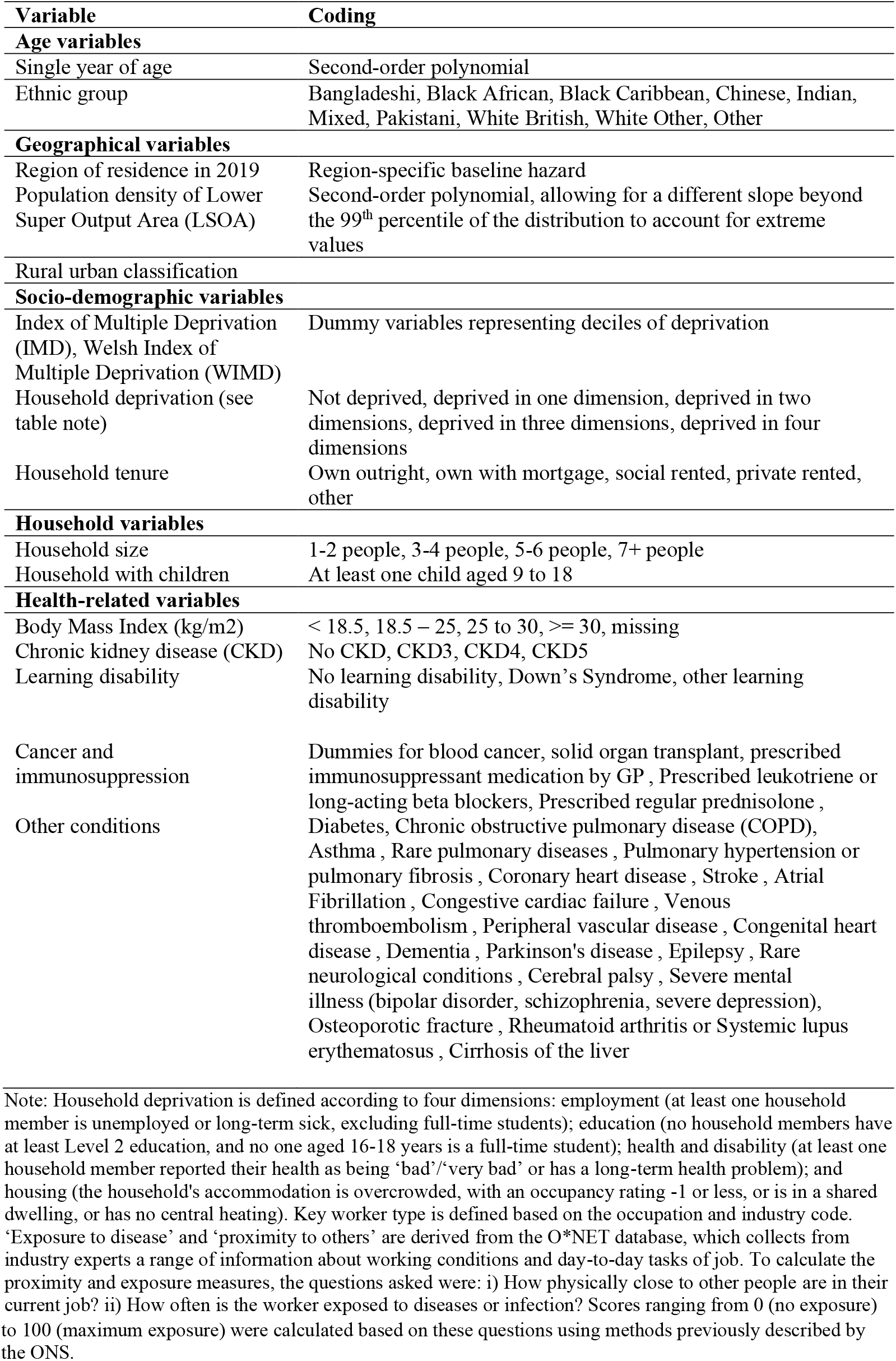
Covariates included in the Cox regression models.

**Supplementary Table S3.**
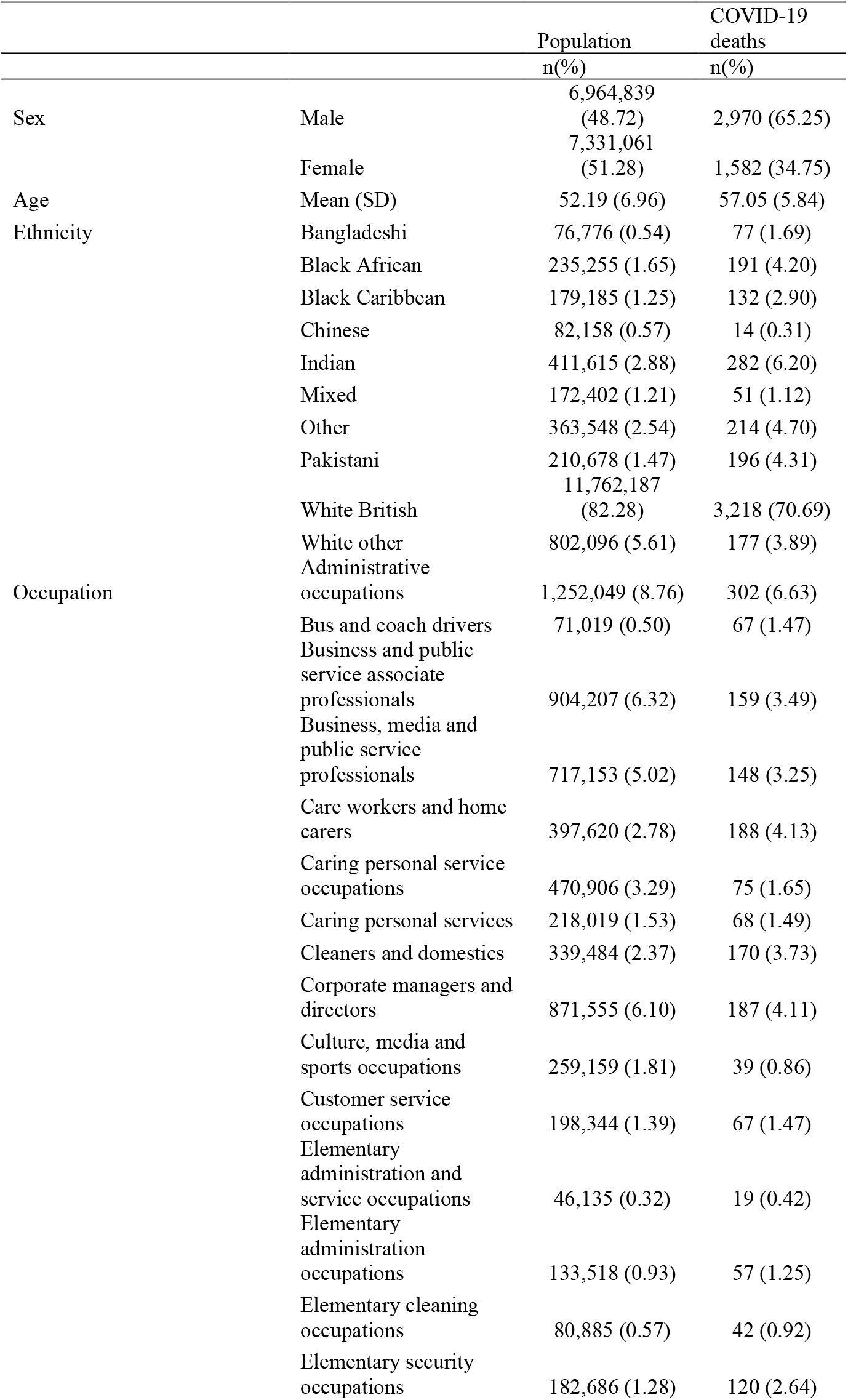
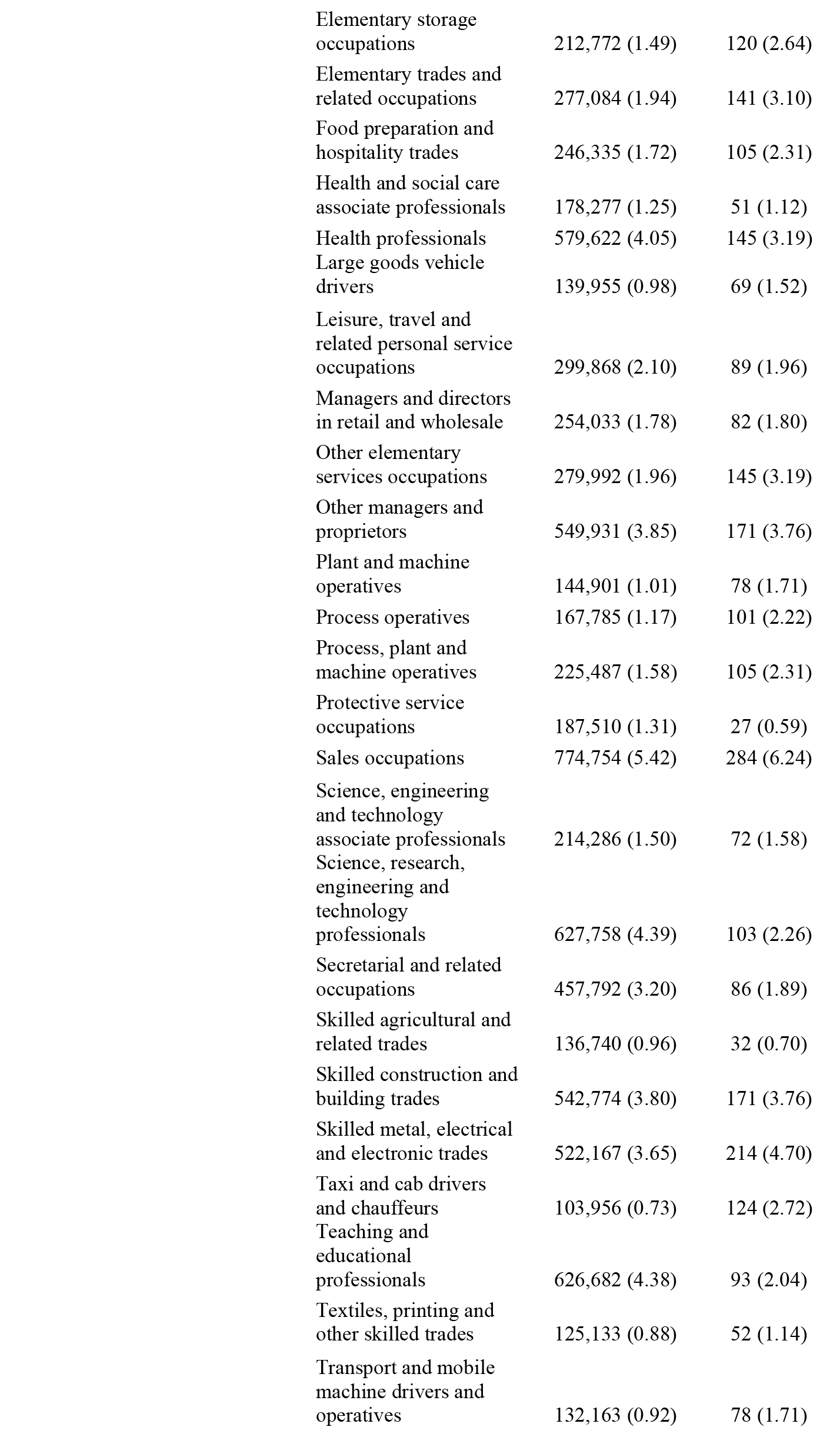
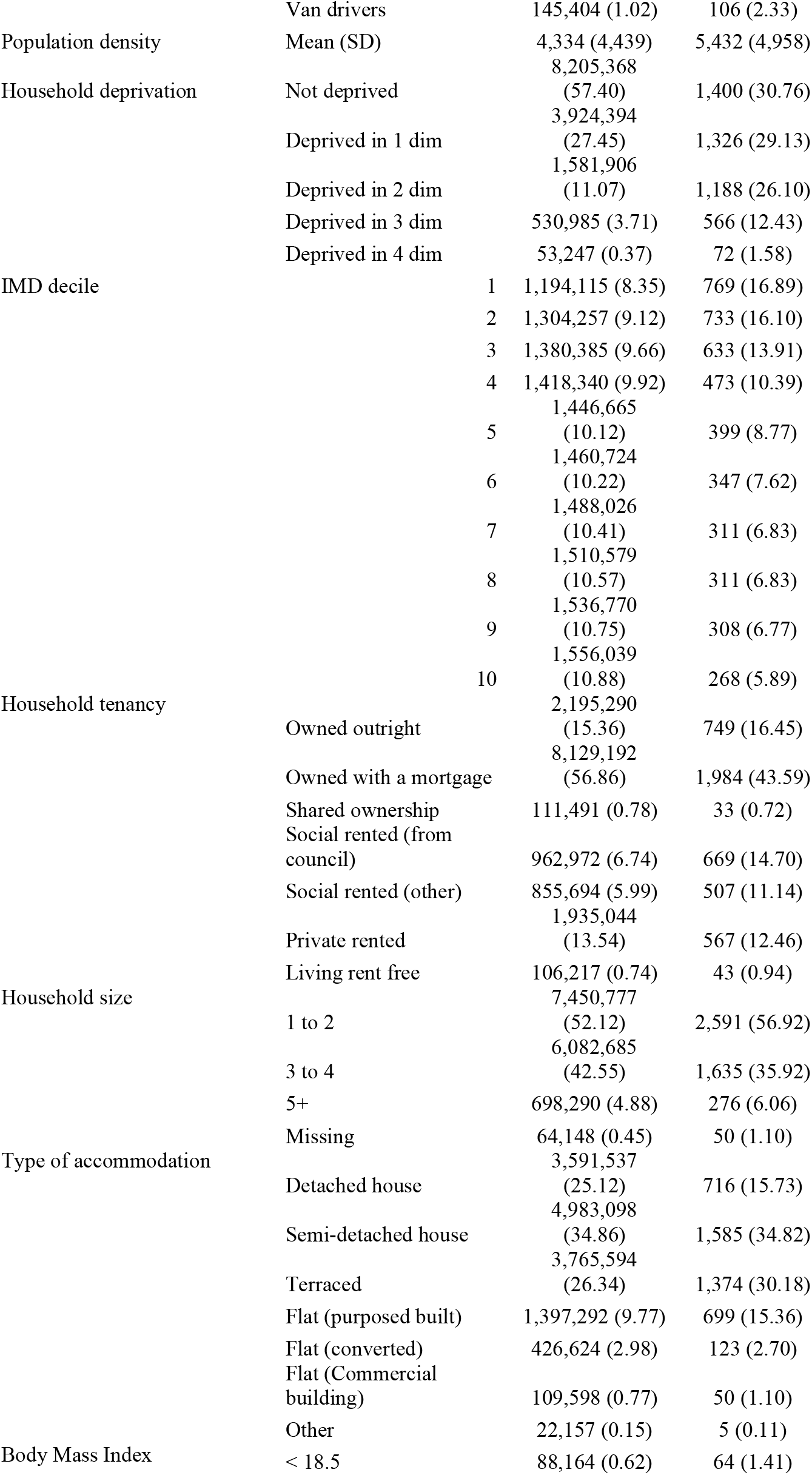
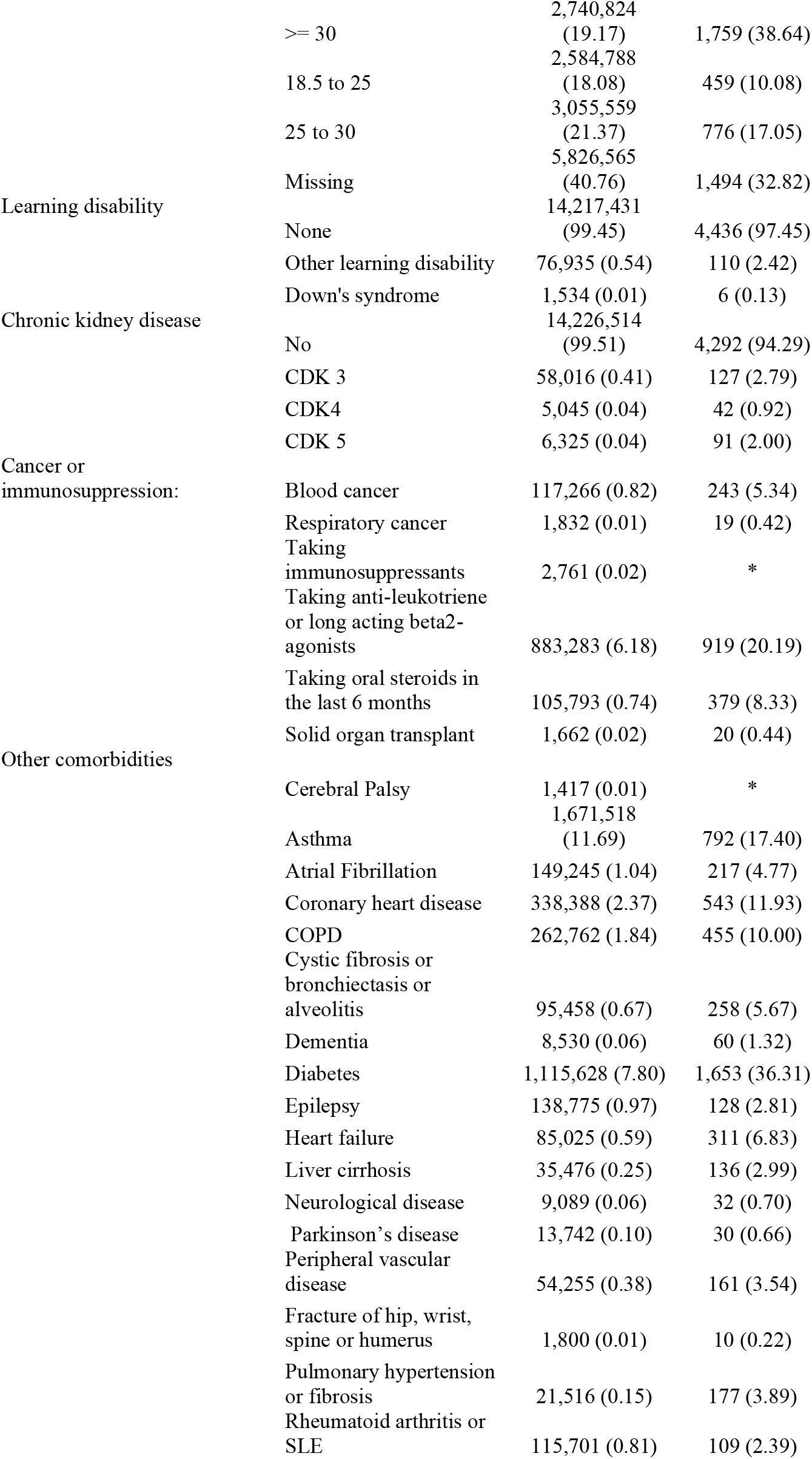
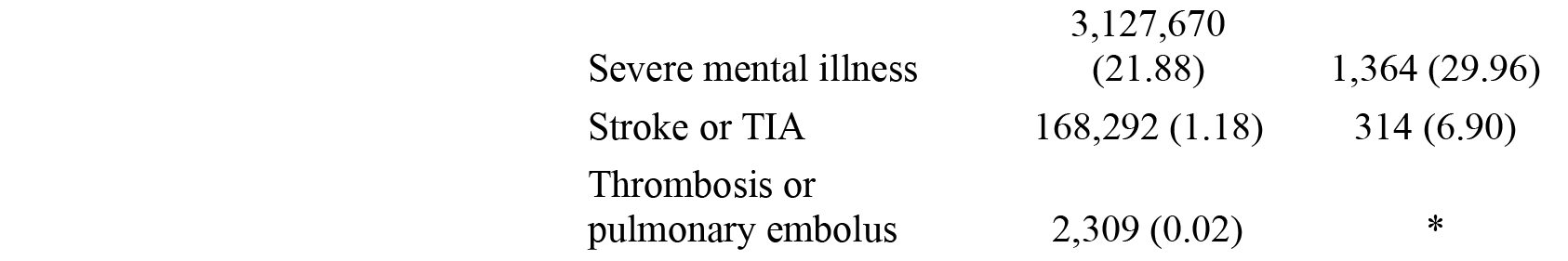
Characteristics of the study population and those who died from a cause related to COVID-19.

**Supplementary Table S4:** Proportion remaining in the same major group occupation in 2011 and 2019, by major group occupation and sex.

| Major occupation group | Men | Women |
| --- | --- | --- |
| Managers, directors and senior officials | 72.1 | 60.8 |
| Professional occupations | 78.9 | 73.9 |
| Associate professional and technical occupations | 68.6 | 65.8 |
| Administrative and secretarial occupations | 63.0 | 69.4 |
| Skilled trades occupations | 74.5 | 62.1 |
| Caring, leisure and other service occupations | 56.1 | 74.4 |
| Sales and customer service occupations | 42.1 | 61.4 |
| Process, plant and machine operatives | 76.2 | 57.9 |
| Elementary occupations | 77.0 | 68.3 |

Source: Understanding Society [ref]

**Supplementary Table S5.** Hazard ratios for COVID-19 related death for men aged 40 to 64, compared to corporate managers and directors.

|  | Model 1 | Model 2 | Model 3 | Model 4 | Model 5 |
| --- | --- | --- | --- | --- | --- |
| Taxi and cab drivers and chauffeurs | 4.62 [3.64 - 5.87] | 3.44 [2.70 - 4.37] | 1.82 [1.41 - 2.34] | 1.57 [1.21 - 2.02] | 1.49 [1.15 - 1.92] |
| Other elementary occupations | 4.22 [3.23 - 5.52] | 3.34 [2.55 - 4.37] | 2.11 [1.60 - 2.79] | 1.55 [1.17 - 2.05] | 1.65 [1.25 - 2.18] |
| Care workers and home carers | 3.89 [2.39 - 6.34] | 3.29 [2.02 - 5.36] | 2.67 [1.64 - 4.36] | 2.13 [1.30 - 3.48] | 2.31 [1.42 - 3.77] |
| Secretarial and related occupations | 3.75 [2.75 - 5.12] | 3.24 [2.37 - 4.42] | 2.36 [1.72 - 3.22] | 1.77 [1.29 - 2.43] | 1.57 [1.14 - 2.15] |
| Elementary security occupations | 3.67 [2.82 - 4.79] | 2.90 [2.22 - 3.79] | 2.00 [1.52 - 2.62] | 1.53 [1.16 - 2.01] | 1.21 [0.92 - 1.59] |
| Bus and coach drivers | 3.50 [2.61 - 4.70] | 2.75 [2.05 - 3.70] | 1.96 [1.45 - 2.65] | 1.54 [1.14 - 2.08] | 1.18 [0.86 - 1.61] |
| Cleaners and domestics | 3.23 [2.35 - 4.44] | 2.61 [1.89 - 3.59] | 1.69 [1.22 - 2.34] | 1.25 [0.90 - 1.74] | 1.27 [0.92 - 1.77] |
| Customer service occupations | 2.97 [2.06 - 4.29] | 2.51 [1.74 - 3.63] | 2.14 [1.48 - 3.10] | 1.77 [1.22 - 2.56] | 1.72 [1.19 - 2.49] |
| Caring personal services | 2.72 [2.12 - 3.50] | 2.33 [1.81 - 3.00] | 1.74 [1.35 - 2.25] | 1.36 [1.05 - 1.77] | 1.33 [1.03 - 1.73] |
| Van drivers | 2.66 [1.75 - 4.02] | 2.31 [1.53 - 3.50] | 1.85 [1.22 - 2.81] | 1.52 [1.00 - 2.31] | 1.37 [0.90 - 2.08] |
| Process operatives | 2.55 [1.94 - 3.36] | 2.23 [1.69 - 2.94] | 1.54 [1.16 - 2.04] | 1.20 [0.91 - 1.60] | 1.21 [0.91 - 1.61] |
| Food preparation and hospitality trades | 2.48 [1.87 - 3.27] | 2.00 [1.52 - 2.65] | 1.23 [0.92 - 1.65] | 0.99 [0.74 - 1.32] | 0.96 [0.71 - 1.28] |
| Sales occupations | 2.46 [1.92 - 3.15] | 2.13 [1.66 - 2.73] | 1.58 [1.23 - 2.04] | 1.19 [0.92 - 1.54] | 1.22 [0.94 - 1.58] |
| Elementary storage occupations | 2.44 [1.90 - 3.12] | 2.06 [1.61 - 2.63] | 1.46 [1.13 - 1.87] | 1.18 [0.92 - 1.52] | 1.17 [0.91 - 1.51] |
| Health and social care associate professionals | 2.27 [1.49 - 3.46] | 1.91 [1.25 - 2.92] | 1.78 [1.16 - 2.71] | 1.49 [0.98 - 2.28] | 1.33 [0.87 - 2.04] |
| Mobile machine and other drivers | 2.27 [1.72 - 2.99] | 2.03 [1.54 - 2.68] | 1.61 [1.22 - 2.13] | 1.28 [0.97 - 1.70] | 1.27 [0.95 - 1.68] |
| Leisure, travel and related personal service occupations | 2.17 [1.55 - 3.04] | 1.79 [1.28 - 2.52] | 1.44 [1.02 - 2.02] | 1.18 [0.84 - 1.66] | 1.23 [0.87 - 1.73] |
| Health professionals | 2.12 [1.58 - 2.84] | 1.93 [1.44 - 2.59] | 1.68 [1.25 - 2.27] | 1.63 [1.21 - 2.20] | 1.65 [1.23 - 2.23] |
| Elementary trades and related occupations | 2.05 [1.59 - 2.64] | 1.85 [1.44 - 2.39] | 1.27 [0.98 - 1.65] | 0.89 [0.69 - 1.16] | 0.93 [0.72 - 1.22] |
| Administrative occupations | 2.00 [1.45 - 2.76] | 1.68 [1.22 - 2.33] | 1.31 [0.94 - 1.81] | 1.09 [0.79 - 1.52] | 1.18 [0.85 - 1.65] |
| Elementary administration occupations | 1.97 [1.57 - 2.48] | 1.69 [1.34 - 2.13] | 1.53 [1.21 - 1.93] | 1.32 [1.05 - 1.67] | 1.33 [1.05 - 1.68] |
| Textiles, printing and other (excl. food prep and hospitality) skilled trades | 1.90 [1.28 - 2.82] | 1.65 [1.11 - 2.45] | 1.22 [0.82 - 1.82] | 0.93 [0.62 - 1.39] | 1.00 [0.67 - 1.49] |
| Elementary cleaning occupations excl. cleaners and domestics | 1.90 [1.34 - 2.67] | 1.68 [1.19 - 2.37] | 1.33 [0.94 - 1.89] | 1.11 [0.78 - 1.57] | 1.11 [0.78 - 1.58] |
| Assemblers and construction operatives | 1.83 [1.39 - 2.41] | 1.64 [1.25 - 2.17] | 1.27 [0.96 - 1.68] | 1.01 [0.76 - 1.34] | 1.01 [0.76 - 1.33] |
| Plant and machine operatives | 1.75 [1.30 - 2.35] | 1.59 [1.18 - 2.13] | 1.24 [0.92 - 1.68] | 0.97 [0.72 - 1.32] | 0.90 [0.67 - 1.22] |
| Large goods vehicle drivers | 1.75 [1.30 - 2.35] | 1.65 [1.23 - 2.23] | 1.56 [1.16 - 2.11] | 1.39 [1.03 - 1.87] | 1.47 [1.09 - 1.98] |
| Managers and directors in retail and wholesale | 1.69 [1.27 - 2.24] | 1.63 [1.23 - 2.17] | 1.33 [1.00 - 1.78] | 1.13 [0.85 - 1.52] | 1.07 [0.80 - 1.42] |
| Science, engineering and technology associate professionals | 1.67 [1.24 - 2.25] | 1.62 [1.20 - 2.19] | 1.35 [1.00 - 1.82] | 1.32 [0.98 - 1.78] | 1.29 [0.96 - 1.75] |
| Skilled metal, electrical and electronic trades | 1.54 [1.25 - 1.90] | 1.44 [1.17 - 1.77] | 1.27 [1.03 - 1.58] | 1.09 [0.88 - 1.35] | 1.10 [0.89 - 1.36] |
| Other managers and proprietors | 1.45 [1.15 - 1.85] | 1.40 [1.10 - 1.77] | 1.14 [0.89 - 1.44] | 1.06 [0.83 - 1.35] | 1.05 [0.82 - 1.33] |
| Caring personal service occupations excl. care workers and home carers | 1.36 [0.70 - 2.67] | 1.24 [0.63 - 2.43] | 1.11 [0.56 - 2.17] | 0.91 [0.46 - 1.78] | 0.84 [0.43 - 1.66] |
| Skilled construction and building trades | 1.22 [0.98 - 1.52] | 1.14 [0.92 - 1.42] | 0.97 [0.78 - 1.22] | 0.82 [0.66 - 1.03] | 0.86 [0.69 - 1.08] |
| Business, media and public service professionals | 1.05 [0.82 - 1.35] | 0.96 [0.75 - 1.24] | 1.06 [0.82 - 1.36] | 1.02 [0.79 - 1.32] | 1.00 [0.77 - 1.29] |
| Culture, media and sports occupations | 1.01 [0.68 - 1.49] | 0.86 [0.58 - 1.27] | 0.92 [0.62 - 1.36] | 0.82 [0.56 - 1.22] | 0.90 [0.61 - 1.33] |
| Business and public service associate professionals | 1.01 [0.79 - 1.29] | 0.97 [0.76 - 1.24] | 0.98 [0.76 - 1.25] | 0.95 [0.74 - 1.21] | 0.94 [0.74 - 1.20] |
| Teaching and educational professionals | 0.99 [0.71 - 1.38] | 0.90 [0.65 - 1.25] | 1.03 [0.73 - 1.44] | 0.96 [0.69 - 1.35] | 1.05 [0.75 - 1.47] |
| Skilled agricultural and related trades | 0.91 [0.61 - 1.36] | 1.14 [0.76 - 1.70] | 0.94 [0.63 - 1.41] | 0.77 [0.52 - 1.16] | 0.85 [0.56 - 1.28] |
| Science, research, engineering and technology professionals | 0.81 [0.62 - 1.05] | 0.79 [0.61 - 1.02] | 0.84 [0.65 - 1.09] | 0.80 [0.62 - 1.04] | 0.82 [0.63 - 1.06] |
| Protective service occupations | 0.74 [0.48 - 1.13] | 0.74 [0.48 - 1.13] | 0.71 [0.47 - 1.09] | 0.68 [0.45 - 1.04] | 0.65 [0.43 - 1.00] |

**Supplementary Table S6.** Hazard ratios for COVID-19 related death for women aged 40 to 64, compared to corporate managers and directors.

|  | Model 1 | Model 2 | Model 3 | Model 4 | Model 5 |
| --- | --- | --- | --- | --- | --- |
| Taxi and cab drivers and chauffeurs | 6.39 [2.49 - 16.39] | 6.14 [2.39 - 15.76] | 4.22 [1.64 - 10.88] | 2.89 [1.12 - 7.47] | 2.77 [1.07 - 7.16] |
| Plant and machine operatives | 5.54 [3.04 - 10.10] | 4.40 [2.41 - 8.04] | 2.96 [1.61 - 5.45] | 1.86 [1.01 - 3.44] | 1.66 [0.89 - 3.08] |
| Bus and coach drivers | 4.79 [1.87 - 12.29] | 4.33 [1.69 - 11.13] | 3.18 [1.23 - 8.19] | 2.16 [0.84 - 5.57] | 1.94 [0.75 - 5.01] |
| Elementary cleaning occupations excl. cleaners and domestics | 4.29 [2.25 - 8.18] | 3.73 [1.95 - 7.11] | 2.57 [1.34 - 4.94] | 1.75 [0.91 - 3.37] | 1.98 [1.03 - 3.81] |
| Process operatives | 3.19 [1.90 - 5.35] | 2.72 [1.62 - 4.58] | 2.00 [1.18 - 3.38] | 1.34 [0.79 - 2.28] | 1.28 [0.75 - 2.17] |
| Elementary trades and related occupations | 3.12 [1.97 - 4.95] | 2.72 [1.71 - 4.31] | 1.88 [1.18 - 3.02] | 1.20 [0.75 - 1.92] | 1.21 [0.75 - 1.94] |
| Elementary security occupations | 3.03 [1.88 - 4.87] | 2.66 [1.65 - 4.29] | 1.92 [1.19 - 3.12] | 1.57 [0.97 - 2.55] | 1.36 [0.83 - 2.21] |
| Care workers and home carers | 2.98 [2.02 - 4.38] | 2.67 [1.81 - 3.93] | 2.12 [1.43 - 3.14] | 1.47 [0.99 - 2.18] | 1.29 [0.87 - 1.92] |
| Cleaners and domestics | 2.89 [1.95 - 4.27] | 2.54 [1.72 - 3.76] | 1.74 [1.17 - 2.61] | 1.24 [0.83 - 1.86] | 1.24 [0.83 - 1.85] |
| Elementary storage occupations | 2.81 [1.50 - 5.26] | 2.43 [1.29 - 4.55] | 1.79 [0.95 - 3.37] | 1.31 [0.70 - 2.48] | 1.58 [0.84 - 2.98] |
| Other elementary occupations | 2.80 [1.86 - 4.21] | 2.52 [1.67 - 3.79] | 1.81 [1.19 - 2.75] | 1.28 [0.84 - 1.95] | 1.26 [0.83 - 1.92] |
| Assemblers and construction operatives | 2.62 [1.59 - 4.34] | 2.18 [1.32 - 3.62] | 1.49 [0.89 - 2.49] | 1.02 [0.61 - 1.70] | 1.03 [0.61 - 1.73] |
| Van drivers | 2.58 [1.01 - 6.63] | 2.50 [0.97 - 6.41] | 1.77 [0.69 - 4.55] | 1.24 [0.48 - 3.20] | 1.43 [0.55 - 3.69] |
| Sales occupations | 2.37 [1.63 - 3.46] | 2.15 [1.48 - 3.14] | 1.57 [1.07 - 2.30] | 1.22 [0.83 - 1.79] | 1.13 [0.77 - 1.67] |
| Mobile machine and other drivers | 2.33 [0.71 - 7.60] | 2.19 [0.67 - 7.14] | 1.90 [0.58 - 6.20] | 1.47 [0.45 - 4.81] | 1.31 [0.40 - 4.31] |
| Skilled metal, electrical and electronic trades | 2.15 [0.76 - 6.09] | 1.86 [0.66 - 5.27] | 1.39 [0.49 - 3.95] | 0.98 [0.34 - 2.77] | 0.88 [0.31 - 2.51] |
| Food preparation and hospitality trades | 2.00 [1.23 - 3.27] | 1.83 [1.12 - 2.98] | 1.46 [0.89 - 2.40] | 1.11 [0.68 - 1.83] | 0.99 [0.60 - 1.63] |
| Customer service occupations | 1.96 [1.20 - 3.21] | 1.77 [1.08 - 2.88] | 1.45 [0.88 - 2.37] | 1.21 [0.74 - 1.98] | 1.14 [0.70 - 1.87] |
| Managers and directors in retail and wholesale | 1.88 [1.10 - 3.22] | 1.80 [1.05 - 3.07] | 1.47 [0.86 - 2.52] | 1.26 [0.74 - 2.16] | 1.37 [0.80 - 2.35] |
| Textiles, printing and other (excl. food prep and hospitality) skilled trades | 1.84 [0.93 - 3.65] | 1.73 [0.87 - 3.43] | 1.29 [0.65 - 2.58] | 0.94 [0.47 - 1.87] | 1.03 [0.52 - 2.06] |
| Caring personal services | 1.82 [1.15 - 2.88] | 1.65 [1.04 - 2.62] | 1.38 [0.87 - 2.20] | 1.10 [0.69 - 1.76] | 1.00 [0.63 - 1.59] |
| Leisure, travel and related personal service occupations | 1.74 [1.11 - 2.73] | 1.65 [1.05 - 2.59] | 1.35 [0.85 - 2.13] | 1.12 [0.71 - 1.77] | 1.25 [0.79 - 1.97] |
| Science, engineering and technology associate professionals | 1.59 [0.82 - 3.09] | 1.54 [0.79 - 2.98] | 1.41 [0.73 - 2.75] | 1.27 [0.65 - 2.47] | 1.32 [0.68 - 2.58] |
| Other managers and proprietors | 1.59 [1.02 - 2.47] | 1.59 [1.02 - 2.47] | 1.45 [0.93 - 2.26] | 1.26 [0.81 - 1.97] | 1.21 [0.78 - 1.89] |
| Health and social care associate professionals | 1.57 [0.94 - 2.63] | 1.44 [0.86 - 2.42] | 1.40 [0.83 - 2.35] | 1.17 [0.70 - 1.97] | 1.21 [0.72 - 2.04] |
| Elementary administration occupations | 1.54 [0.73 - 3.23] | 1.41 [0.67 - 2.95] | 1.05 [0.50 - 2.21] | 0.80 [0.38 - 1.68] | 0.80 [0.38 - 1.69] |
| Health professionals | 1.38 [0.92 - 2.08] | 1.31 [0.87 - 1.97] | 1.53 [1.01 - 2.31] | 1.37 [0.91 - 2.08] | 1.30 [0.86 - 1.97] |
| Administrative occupations | 1.36 [0.93 - 1.99] | 1.27 [0.87 - 1.86] | 1.10 [0.75 - 1.61] | 1.00 [0.68 - 1.46] | 1.01 [0.69 - 1.49] |
| Skilled construction and building trades | 1.34 [0.32 - 5.58] | 1.27 [0.30 - 5.29] | 1.02 [0.24 - 4.27] | 0.78 [0.19 - 3.26] | 0.87 [0.21 - 3.62] |
| Caring personal service occupations excl. care workers and home carers | 1.28 [0.84 - 1.95] | 1.22 [0.80 - 1.86] | 1.08 [0.71 - 1.65] | 0.96 [0.63 - 1.47] | 0.99 [0.65 - 1.52] |
| Business, media and public service professionals | 1.28 [0.81 - 2.01] | 1.24 [0.79 - 1.94] | 1.40 [0.89 - 2.21] | 1.35 [0.86 - 2.12] | 1.35 [0.86 - 2.12] |
| Skilled agricultural and related trades | 1.23 [0.44 - 3.49] | 1.60 [0.56 - 4.54] | 1.39 [0.49 - 3.94] | 1.19 [0.42 - 3.37] | 1.13 [0.39 - 3.23] |
| Secretarial and related occupations | 1.19 [0.78 - 1.82] | 1.16 [0.76 - 1.77] | 0.98 [0.64 - 1.49] | 0.91 [0.59 - 1.39] | 0.94 [0.62 - 1.44] |
| Business and public service associate professionals | 1.08 [0.70 - 1.69] | 1.05 [0.68 - 1.64] | 1.02 [0.66 - 1.60] | 0.97 [0.62 - 1.52] | 1.01 [0.65 - 1.58] |
| Teaching and educational professionals | 0.88 [0.56 - 1.37] | 0.86 [0.55 - 1.34] | 1.01 [0.64 - 1.59] | 0.98 [0.63 - 1.54] | 1.06 [0.67 - 1.66] |
| Science, research, engineering and technology professionals | 0.80 [0.41 - 1.60] | 0.79 [0.40 - 1.57] | 0.90 [0.45 - 1.78] | 0.87 [0.44 - 1.73] | 0.93 [0.47 - 1.84] |
| Culture, media and sports occupations | 0.67 [0.32 - 1.40] | 0.67 [0.32 - 1.39] | 0.72 [0.34 - 1.51] | 0.68 [0.32 - 1.42] | 0.70 [0.33 - 1.50] |
| Protective service occupations | 0.45 [0.11 - 1.88] | 0.45 [0.11 - 1.86] | 0.39 [0.09 - 1.63] | 0.37 [0.09 - 1.54] | 0.38 [0.09 - 1.61] |

**Supplementary Table S7.** Hazard ratios for COVID-19 related death for adults aged 40 to 64, compared to all other occupations, by sex.

| Occupation | Men |  | Women |  |
| --- | --- | --- | --- | --- |
|  | Age adjusted | Fully adjusted | Age adjusted | Fully adjusted |
| Taxi and cab drivers and chauffeurs | 2.81 [2.34 - 3.38] | 1.32 [1.09 - 1.60] | 3.62 [1.5 - 8.7] | 2.34 [0.97 - 5.64] |
| Other elementary occupations | 2.55 [2.04 - 3.18] | 1.47 [1.17 - 1.84] | 1.67 [1.34 - 2.09] | 1.09 [0.87 - 1.36] |
| Secretarial and related occupations | 2.32 [1.46 - 3.68] | 2.07 [1.30 - 3.29] | 0.64 [0.5 - 0.82] | 0.83 [0.65 - 1.06] |
| Care workers and home carers | 2.25 [1.72 - 2.94] | 1.39 [1.05 - 1.82] | 1.84 [1.54 - 2.19] | 1.14 [0.95 - 1.36] |
| Elementary security occupations | 2.21 [1.78 - 2.75] | 1.06 [0.85 - 1.32] | 1.81 [1.3 - 2.52] | 1.14 [0.81 - 1.61] |
| Bus and coach drivers | 2.10 [1.63 - 2.70] | 1.04 [0.79 - 1.36] | 2.83 [1.18 - 6.81] | 1.57 [0.65 - 3.8] |
| Cleaners and domestics | 1.93 [1.46 - 2.55] | 1.12 [0.84 - 1.49] | 1.82 [1.51 - 2.19] | 1.12 [0.92 - 1.36] |
| Customer service occupations | 1.77 [1.27 - 2.47] | 1.54 [1.10 - 2.15] | 1.12 [0.79 - 1.59] | 1.04 [0.73 - 1.47] |
| Van drivers | 1.64 [1.34 - 2.00] | 1.20 [0.98 - 1.46] | 1.42 [0.59 - 3.42] | 1.32 [0.55 - 3.17] |
| Caring personal services | 1.58 [1.07 - 2.32] | 1.21 [0.82 - 1.79] | 1.05 [0.77 - 1.42] | 0.9 [0.66 - 1.23] |
| Process operatives | 1.53 [1.21 - 1.92] | 1.08 [0.86 - 1.36] | 2.06 [1.4 - 3.03] | 1.05 [0.71 - 1.55] |
| Food preparation and hospitality trades | 1.48 [1.17 - 1.87] | 0.82 [0.64 - 1.04] | 1.21 [0.85 - 1.71] | 0.9 [0.63 - 1.28] |
| Elementary storage occupations | 1.48 [1.22 - 1.79] | 1.09 [0.90 - 1.33] | 1.66 [0.98 - 2.82] | 1.33 [0.78 - 2.25] |
| Sales occupations | 1.46 [1.21 - 1.77] | 1.03 [0.85 - 1.25] | 1.38 [1.18 - 1.62] | 0.98 [0.84 - 1.16] |
| Mobile machine and other drivers | 1.35 [1.08 - 1.70] | 1.14 [0.91 - 1.44] | 1.34 [0.43 - 4.17] | 1.29 [0.41 - 4] |
| Health and social care associate professionals | 1.35 [0.91 - 2.00] | 1.19 [0.80 - 1.77] | 0.91 [0.62 - 1.34] | 1.04 [0.71 - 1.54] |
| Leisure, travel and related personal service occupations | 1.29 [0.96 - 1.75] | 1.10 [0.81 - 1.48] | 1 [0.75 - 1.35] | 1.06 [0.79 - 1.43] |
| Health professionals | 1.26 [0.99 - 1.62] | 1.54 [1.19 - 1.99] | 0.82 [0.66 - 1.03] | 1.23 [0.96 - 1.57] |
| Elementary trades and related occupations | 1.22 [1.00 - 1.49] | 0.82 [0.67 - 1.01] | 2.03 [1.49 - 2.75] | 1.04 [0.76 - 1.42] |
| Elementary administration occupations | 1.19 [0.89 - 1.58] | 1.05 [0.79 - 1.40] | 0.91 [0.47 - 1.75] | 0.7 [0.36 - 1.36] |
| Administrative occupations | 1.18 [0.99 - 1.40] | 1.20 [1.01 - 1.43] | 0.73 [0.62 - 0.86] | 0.84 [0.71 - 0.99] |
| Elementary cleaning occupations excl. cleaners and domestics | 1.13 [0.78 - 1.63] | 0.89 [0.62 - 1.28] | 2.69 [1.56 - 4.65] | 1.8 [1.04 - 3.12] |
| Textiles, printing and other (excl. food prep and hospitality) skilled trades | 1.13 [0.83 - 1.53] | 1.00 [0.73 - 1.36] | 1.05 [0.58 - 1.9] | 0.85 [0.47 - 1.54] |
| Assemblers and construction operatives | 1.09 [0.87 - 1.37] | 0.90 [0.71 - 1.13] | 1.67 [1.15 - 2.41] | 0.82 [0.56 - 1.2] |
| Plant and machine operatives | 1.04 [0.81 - 1.33] | 0.80 [0.62 - 1.04] | 3.42 [2.09 - 5.6] | 1.67 [1.02 - 2.75] |
| Science, engineering and technology associate professionals | 1.04 [0.80 - 1.34] | 1.35 [1.04 - 1.74] | 0.91 [0.51 - 1.6] | 1.12 [0.63 - 1.97] |
| Large goods vehicle drivers | 1.00 [0.79 - 1.27] | 0.96 [0.76 - 1.22] | - | - |
| Managers and directors in retail and wholesale | 0.99 [0.76 - 1.28] | 1.17 [0.90 - 1.52] | 1.05 [0.7 - 1.59] | 1.25 [0.82 - 1.88] |
| Skilled metal, electrical and electronic trades | 0.91 [0.79 - 1.05] | 1.01 [0.87 - 1.16] | 1.28 [0.48 - 3.41] | 0.73 [0.27 - 1.95] |
| Other managers and proprietors | 0.86 [0.71 - 1.03] | 0.94 [0.78 - 1.13] | 0.92 [0.7 - 1.21] | 1.14 [0.86 - 1.5] |
| Caring personal service occupations excl. care workers and home carers | 0.81 [0.42 - 1.55] | 0.75 [0.39 - 1.44] | 0.7 [0.55 - 0.89] | 0.85 [0.66 - 1.09] |
| Skilled construction and building trades | 0.71 [0.60 - 0.83] | 0.77 [0.66 - 0.90] | 0.77 [0.19 - 3.08] | 0.8 [0.2 - 3.2] |
| Business, media and public service professionals | 0.61 [0.50 - 0.74] | 0.91 [0.74 - 1.12] | 0.75 [0.56 - 1.01] | 1.25 [0.93 - 1.7] |
| Culture, media and sports occupations | 0.60 [0.42 - 0.85] | 0.82 [0.57 - 1.17] | 0.39 [0.2 - 0.75] | 0.68 [0.35 - 1.31] |
| Business and public service associate professionals | 0.58 [0.48 - 0.70] | 0.86 [0.70 - 1.04] | 0.6 [0.45 - 0.79] | 0.89 [0.67 - 1.18] |
| Teaching and educational professionals | 0.58 [0.43 - 0.78] | 0.96 [0.71 - 1.30] | 0.48 [0.36 - 0.64] | 0.93 [0.69 - 1.26] |
| Corporate managers and directors excl. those in retail and wholetrade | 0.57 [0.49 - 0.67] | 0.92 [0.78 - 1.08] | 0.55 [0.39 - 0.78] | 0.86 [0.61 - 1.23] |
| Skilled agricultural and related trades | 0.53 [0.37 - 0.77] | 0.77 [0.53 - 1.13] | 0.66 [0.25 - 1.76] | 0.99 [0.36 - 2.7] |
| Science, research, engineering and technology professionals | 0.46 [0.38 - 0.57] | 0.73 [0.59 - 0.91] | 0.48 [0.26 - 0.87] | 0.84 [0.46 - 1.53] |
| Protective service occupations | 0.43 [0.29 - 0.64] | 0.59 [0.40 - 0.88] | 0.24 [0.06 - 0.97] | 0.37 [0.09 - 1.46] |

Note: each line represents a separate regression in which each occupational group is compared with all other workers. Fully adjusted Cox regression models include geographical factors (region, population density, urban/rural classification), ethnicity, socio-economic characteristics (IMD decile, household deprivation, educational attainment, social grade, household tenancy, type of accommodation, household size, multigenerational household, household with children), health (Body Mass Index, Chronic kidney disease (CKD), Learning disability, Cancer and immunosuppression, other conditions) See Supplementary Tables A1 for more details.

**Supplementary Table S7.** Hazard ratios for COVID-19 related death for adults aged 40 to 64, Major occupational group, stratified by sex.

| Occupation | Men |  | Women |  |
| --- | --- | --- | --- | --- |
|  | Age adjusted | Fully adjusted | Age adjusted | Fully adjusted |
| skilled trades occupations | 2.15 [1.87 - 2.47] | 1.13 [0.98 - 1.32] | 2.07 [1.66 - 2.58] | 1.08 [0.86 - 1.36] |
| professional occupations | 2.12 [1.74 - 2.58] | 1.17 [0.96 - 1.43] | 1.68 [1.33 - 2.12] | 1.02 [0.80 - 1.29] |
| process, plant and machine operatives | 2.12 [1.73 - 2.59] | 1.19 [0.96 - 1.46] | 1.41 [1.13 - 1.77] | 1.01 [0.81 - 1.27] |
| sales and customer service occupations | 2.01 [1.76 - 2.30] | 1.16 [1.01 - 1.34] | 2.39 [1.80 - 3.17] | 1.16 [0.87 - 1.56] |
| caring, leisure and other service occupations | 1.69 [1.40 - 2.05] | 1.32 [1.09 - 1.61] | 0.94 [0.75 - 1.18] | 0.87 [0.69 - 1.10] |
| elementary occupations | 1.19 [1.04 - 1.37] | 0.96 [0.83 - 1.10] | 1.32 [0.95 - 1.84] | 0.82 [0.59 - 1.15] |
| associate professional and technical occupations | 0.94 [0.79 - 1.10] | 0.99 [0.84 - 1.17] | 0.81 [0.62 - 1.07] | 0.90 [0.68 - 1.18] |
| administrative and secretarial occupations | 0.86 [0.74 - 1.01] | 0.95 [0.80 - 1.12] | 0.82 [0.65 - 1.05] | 1.05 [0.81 - 1.35] |

Note: Reference – Managers and directors. Fully adjusted Cox regression models include geographical factors (region, population density, urban/rural classification), ethnicity, socio-economic

